# The curvilinear relationships between grand parity and incidence of hormone-dependent cancers; follow-up of postmenopausal women in the Norwegian 1960 Census

**DOI:** 10.1101/2023.12.13.23299895

**Authors:** Eiliv Lund, Lill-Tove Rasmussen Busund, Lars Holden

**Affiliations:** UiT The Arctic University of Norway, Tromsø, Norway; Department of Medical Biology, UiT The Arctic University of Norway, Tromsø, Norway; Department of Clinical Pathology, University Hospital of North Norway, Tromsø, Norway; Norwegian Computing Center, Oslo, Norway

**Keywords:** parity, pregnancy, breast cancer, ovarian cancer, endometrial cancer, Norwegian Census 1960, cohort design, collinearity

## Abstract

Breast, ovarian and endometrial cancers are named hormone dependent cancers (HDC) because of the effects of endogenous hormones, including parity, on the incidence rates. Here, we will test the hypothesis that each additional child has the same relative preventive effect on the risk of each of the three cancer sites and with similar shapes of the incidence curves over the extended exposure range, 1 - 18 children in postmenopausal women.

The study is based on parity information from the Norwegian 1960 Census for women aged 45-89 years. A total of 385 816 married women answered the question of number of children in present marriage to a civil servant. Follow-up continued until the first of any of the HDC diagnosis, death, or end of follow-up 2005 through linkages based on the unique Norwegian birth number. Included were 16 905 breast cancers, 3 827 ovarian cancers and 3 834 endometrial cancers. Based on person-years (PY), the percentage change in incidence rates of the three cancer sites for each additional child was calculated using a logit regression model including models with higher order terms. A new statistical method for analyses of collinearity between parity and age at first birth has been developed. Age at marriage was used as a proxy for age at first birth.

Parity had strong linear effects on the incidence rates for all three cancer sites (p<2e-16). The percentage decrease for each of the cancer diagnosis for an additional child were for breast cancer 10.5% (95% CI; 9.6-11.4), ovarian cancer 13.2% (11.2-15.3) and endometrial cancer 10.9% (8.9-12.8) with similar curvilinear relationship. Models with higher order terms gave slightly better fit to the data with a stronger protective effect for increasing parity on ovarian cancers, while for breast cancer age became more important. Combining the incidence of all three cancers gave a percentage decrease for each additional child of 11.0% (10.1-11.8). The risk for HDC cancer was reduced with earlier age at marriage (first birth) for women with 1-2 children for breast cancer and ovarian cancer and 1 child for endometrial cancer, but the effect was smaller than one additional child. Age of the women at marriage was not significant for the sum of the three cancers.

The study demonstrated the strong, regular protective effect of each additional child or full-term pregnancy on the incidence rates of the hormone-dependent cancers throughout the exposure range and with similar curvilinear relations. Reduced fertility is a common, strong etiological factor for the three hormone-dependent cancers in postmenopausal women. These findings support a hypothesis that similar immunological changes during each pregnancy could be the biological explanation.

## Introduction

Cancer of the breast, ovary and endometrium have been named the hormone-dependent cancers, HDC, (1, 2) due to the associations with changing levels of endogenous hormones related to menstruation such as age at menarche and menopause, and pregnancy factors like parity, age at first birth and lactation. Parity has been the first and most intensively studied over the last century (3, 4, 5). Only a few studies have been performed with all three cancer sites in the same study yielding directly comparable estimates of either incidence (6, 7, 8) or mortality (9, 10, 11, 12). Some few cohort studies have shown estimates for grand-grand multiparity (GGM), 10 or more children (7, 9). Neither of the studies have described the linear trends for either of the HDC over the extended exposure range up to 18 children, nor compared the curvilinear relationship for each of the HDC separately or in combination. Almost all current large cohorts have a restricted exposure range for parity with lack of women with grand parity, five or more children partly due to the low fertility for decades in most western countries. This is described further in Discussions. In today’s high fertility countries in Africa and Asia large cohorts or linkage studies is difficult to run due to lack of registers and infrastructure

In a previous Norwegian linkage study information on number of children at the 1970 Census were linked to the Cause of Death Register in Norway. Both analyses of each cancer site separately (13, 14, 15) or combined (9) showed regular decrease of mortality rates for increasing parity, but rather few grand-grand multiparous women reduced the statistical power.

To explore the common curvilinear relationship between number of full-term pregnancies or parity and the three HDC, a historical cohort study based on all married women in the 1960 Norwegian Census with follow-up through linkages to the national cancer register has been created. The strong design feature using the 1960 Census is the creation of a unique birth number for all persons alive in Norway at the time of the census (16). This number have made linkage studies feasible. This study will cover women born between 1871 and 1915, a period with many grand-grand parous women. From the 1960 Census forward till the 1970 Census the numbers of married women with 10 children decreased from 5452 to 2386 or 55%. Changing the analyses from mortality to incidence with a prolonged follow-up will increase the statistical power considerable. Based on previous work (9,12,13,14) we aim at testing the biological hypothesis that each additional full-term pregnancy or child, has similar protective effect in all three hormone-dependent cancers with the same overall curvilinear relationship over the extended exposure range of 1-18 children.

## Material and methods

### Material

The 1960 Census in Norway was the source for the creation of the population register of Norway. The information was gathered from every household by civil servants (16). For social reasons the questions of number of children were only posed to married women and only children born in the actual marriage were counted. All persons in Norway alive November 1960 were counted and given a unique birth number. The study population consists of all married women aged 45-89 years at the Norwegian 1960 Census. Women aged 45 years, or more are defined as postmenopausal. The information used for the analysis is number of children, age at marriage and age in 1960 of the women. There are 386 114 unique women in the data set and 385 816 where the number of children in the present marriage is known, see Table 1. For 298 women the number of children was not specified. Of the women 64.7% was aged 45-59, 30.7% 60-74 and 4.6% 75-89. Table 2 shows the average age and the quantiles of age when the women were married as a function of number of children in the present marriage. The age of marriage decreases with the number of children in the marriage. In particular, the age when marrying is much higher for women without children in the present marriage. A number of these women might have children in a previous marriage that was not registered in the census data.

**Table 1.** Number of women in the 1960 Census after parity and age-groups in the current marriage (Statistics Norway). Cell with less than 5 women are truncated due to privacy.

| Age\chi | 0 | 1 | 2 | 3 | 4 | 5 | 6 | 7 | 8 | 9 |
| --- | --- | --- | --- | --- | --- | --- | --- | --- | --- | --- |
| 45-59 | 42758 | 43932 | 57019 | 40098 | 25514 | 15229 | 9408 | 6037 | 3941 | 2499 |
| 60-74 | 20322 | 20719 | 26792 | 19162 | 12178 | 7241 | 4350 | 2870 | 1860 | 1201 |
| 75-89 | 3078 | 3008 | 3968 | 3011 | 1857 | 1081 | 703 | 396 | 291 | 201 |
| Sum | 66158 | 67659 | 87779 | 62271 | 39549 | 23551 | 14461 | 9303 | 6092 | 3901 |
| Age\chi | 10 | 11 | 12 | 13 | 14 | 15-21 | 15 | 16 | 17 | 18-21 |
| 45-59 | 1562 | 826 | 480 | 209 | 94 |  | 42 | 13 | 6 | 11 |
| 60-74 | 776 | 393 | 239 | 104 | 50 | 28 |  |  |  |  |
| 75-89 | 116 | 64 | 37 | 24 | 10 | 8 |  |  |  |  |
| Sum | 2454 | 1283 | 756 | 337 | 154 |  | 64 | 22 | 8 | 14 |

**Table 2.** Age at marriage in the present marriage as function of age at the 1960 Census and number of children in the present marriage; the age at different quantiles.

| Children | 0 | 1 | 2 | 3 | 4 | 5 | 6 | 7 |
| --- | --- | --- | --- | --- | --- | --- | --- | --- |
| Av. Age | 36.5 | 28.8 | 26.8 | 25.8 | 25.0 | 24.3 | 23.9 | 23.3 |
| 10% | 24 | 22 | 21 | 21 | 20 | 20 | 20 | 19 |
| 20% | 27 | 23 | 23 | 22 | 21 | 21 | 21 | 20 |
| 50% | 36 | 28 | 26 | 25 | 24 | 24 | 23 | 23 |
| 80% | 45 | 34 | 31 | 29 | 28 | 27 | 27 | 26 |
| 90% | 49 | 37 | 34 | 32 | 31 | 30 | 29 | 28 |
| Children | 8 | 9 | 10 | 11 | 12 | 13 | 14 | 14+ |
| Av. Age | 22.9 | 22.4 | 22.0 | 21.5 | 21.3 | 20.6 | 20.6 | 20.4 |
| 10% | 19 | 19 | 19 | 19 | 18 | 18 | 18 | 18 |
| 20% | 20 | 20 | 20 | 20 | 19 | 19 | 19 | 18 |
| 50% | 23 | 22 | 22 | 22 | 21 | 21 | 20 | 20 |
| 80% | 26 | 25 | 25 | 24 | 24 | 23 | 22 | 22 |
| 90% | 28 | 27 | 26 | 26 | 25 | 25 | 23 | 23 |

The 1960 Census did not ask women about the age at each birth in contrast to the 1970 Census (9). Due to the discussions around the effect of age at first birth mainly for breast cancer age at marriage is introduced as an approximation. In 1965 the Central Bureau of Statistics in Norway published an analysis showing the close relationship between age at marriage and age at first full-term pregnancy (17). As an example, among women married 1946-1950 aged 18-20 years 81% had a child within 2 years marriage compared to 61% in women aged 25-29 years. Of the 385 816 women with number of children, age at marriage (interval 16,45 years) is known for 319 454 women with at least one child. 66 158 women did not have children in their current marriage.

The effects of collinearity between number of children and age at first and last birth is illustrated in Figure 1 using the 10% and 90% percentiles from Table 2. It is assumed that the first child is born *max(2 exp(-0*.*1(n-1)),1)* after the marriage, which is 2 years for n=1 child and decreases for higher number of children, n. Further, the average interval between the children follows the function *max(4 exp(-0*.*08(n-2)),1*.*5)* which is 4 years for n=2 children and decreases to 1.5 for n=14 children. These functions are based on data for marriages 1946-1960 given by Statistics Norway (17). This gives a 14-year interval for the first birth for women with 1 child, 7 years interval for women with 10 children and 5 years interval for women with 15 children. The uncertainty in the distribution of the age of the mother for the last child is larger than the uncertainty at the time of the first child. While a woman with two children, the normal in Norway today, can have both deliveries early in the twenties or both late in the thirties, a woman with ten children must start early and end late. Hence, there are no women with late start and high parity. The statistical limitations of observed values for both age at first and last birth with increasing parity has not been discussed in statistical analyses of grand multiparous women. A new method is described under Statistical methods.

**Figure 1.**
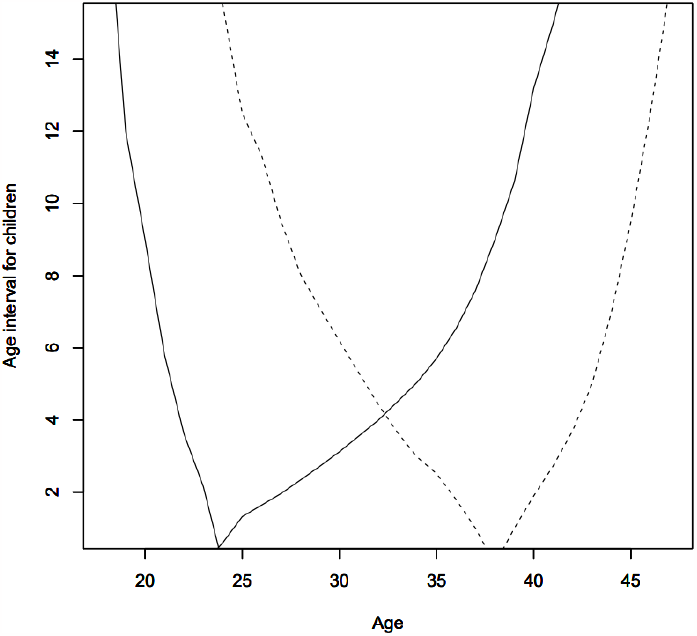
The dependency of age at first and last birth with increasing on parity, based on numbers from Table 2 and estimated intervals between births. The two lines are age at first and last birth for the 10% of women with an early birth and the 10% women with early last birth. Showing decreasing age with increasing parity. The dashed lines are the 90% quantile for the first and last child. With 10 children is the 80% interval for the first birth (20,27) years and for the last birth (39,46)

The project has been evaluated and accepted by the Regional Ethical Committee for South-East Norway (number 475656) and approved 12.10.2022.

### Follow-up

Follow-up to the Norwegian Cancer Register and the register of death certificates was based on the unique birth number given all women alive at the 1960 Census. Follow-up was terminated at 31.12.2005. The main analyses will use cancer incidence diagnoses for the three hormone-dependent cancer sites: breast, endometrium, and ovary. In addition, information on cervical cancer was included as a non-hormonal comparison since the major causal factor is human papilloma virus (18). The Cancer Registry of Norway started in 1953 and had reached good national coverage at start of follow-up. The international codes for diseases ICD7, ICD8, ICD9 and ICD 10 have been transformed to a common version of ICD10 by the register. The linkage to the Norwegian Cancer Registry gave a total of 29 730 diagnosis. Sarcomas (n=62) were excluded together with in-situ (n=272). For women with two or three HDC diagnosis or cervical cancer only the first diagnosis was included (n=1063). Women with cancer above 89 years (n=1047) and with unknown parity (n=13) were excluded leaving a study population of 27 273 cases. The study covers 16 905 breast cancer cases, 3 827 ovarian cancers, 3 834 endometrial cancers and 2 707 cervical cancers.

Causes of death were available from Central Bureau of Statistics. Date of death independent of cause were extracted while cause of death was included only for the three hormone-dependent cancers in the period 1960-2005, i.e., when all women in the Cohort is at least 90 years old. No information can be obtained for date of death of the youngest women in the 1960 census that lived longer than 90 years and the very small number of women that emigrated after 1960. Number of women not registered in the Cause of Death registry for 5- and 10-years cohorts is shown in Table 3. The number of women that emigrates is less than 0,2%. For the 742 women that would have been above 100 years in 2005, end of follow-up is set to 1970, 10 years after the census.

**Table 3.** Average age at death according to parity and age at start of follow-up based on information from the Cause of Death Registry and Statistics Norway. Calculated in cells with at least 5 women.

| Age\chi | 0 | 1 | 2 | 3 | 4 | 5 | 6 | 7 | 8 |
| --- | --- | --- | --- | --- | --- | --- | --- | --- | --- |
| **45-59** | 79.9 | 80.3 | 80.5 | 80.4 | 80.3 | 80.1 | 80.1 | 79.8 | 80.0 |
| **60-74** | 81.8 | 82.1 | 82.2 | 82.1 | 82.1 | 82.0 | 82.0 | 81.8 | 81.4 |
| **75-89** | 85.7 | 85.7 | 86.1 | 85.9 | 86.1 | 86.2 | 86.0 | 85.9 | 85.3 |
| **Sum** | 80.6 | 80.9 | 81.2 | 81.3 | 81.5 | 81.6 | 81.8 | 81.8 | 81.8 |
| Age\chi | 9 | 10 | 11 | 12 | 13 | 14 | 15 | 16 | 17 |
| **45-59** | 79.5 | 78.9 | 78.6 | 77.8 | 80.8 | 74.1 | 79.8 |  |  |
| **60-74** | 81.6 | 81.6 | 81.7 | 81.1 | 81.8 | 81.2 | 79.2 | 84.0 | 81.7 |
| **75-89** | 85.9 | 85.3 | 85.4 | 86.4 | 86.2 | 82.8 |  |  |  |
| **Sum** | 81.8 | 81.7 | 81.8 | 80.8 | 82.7 | 79.7 | 81.0 | 82.1 | 83.2 |

**Table 4.**
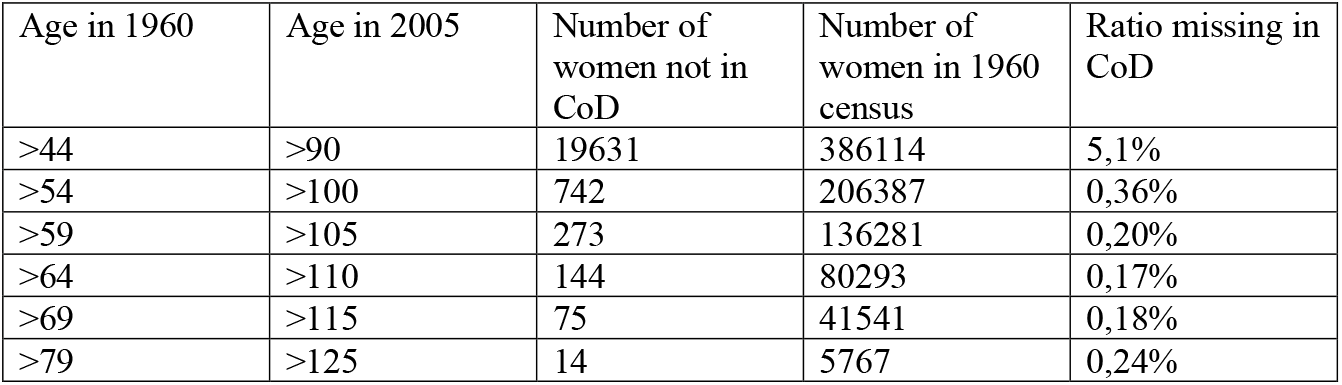
The number of women not found in the Cause of Death (CoD) register for different age cohorts. Includes women with unknow number of children.

| Age in 1960 | Age in 2005 | Number of women not in CoD | Number of women in 1960 census | Ratio missing in CoD |
| --- | --- | --- | --- | --- |
| >44 | >90 | 19631 | 386114 | 5,1% |
| >54 | >100 | 742 | 206387 | 0,36% |
| >59 | >105 | 273 | 136281 | 0,20% |
| >64 | >110 | 144 | 80293 | 0,17% |
| >69 | >115 | 75 | 41541 | 0,18% |
| >79 | >125 | 14 | 5767 | 0,24% |

For each woman the number of person-year is calculated from the age at the 1960 census and until the first of the following: age 90, death or the first cancer diagnosis of the four selected for these analyses.

### Statistical methods

The confidence intervals for the probability per year for the four types of cancers were estimated as a function of number of children. For 11 children and more, the estimate for n children is based on the observations for n, n+1 and n+2 and part of the observations with n-1 children such that the average number of children is n. This implies that the intervals for less than 9 children are independent for each number of children and that there is a slight smoothing of the curves above 8 children due to partly use of the same data. The uncertainty is estimated from the binomial distribution based on the number of observed cancers and the number of years with women with the specified number of children.

The probability for the four cancers was modelled in a logit regression model using number of children and age as covariates. A linear logit model implies that the following three parameter model was estimated for the probability for cancer:

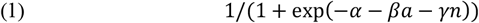

where *a* is the age and *n* is the number of children. This model gives the reduction in probability of cancer

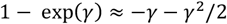

when adding one more child. This is independent of *n*, the number of children. A logit model with higher order terms, i.e. six parameters were included:

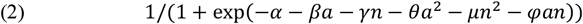

This model gives the reduction in probability of cancer

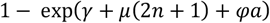

when adding one more child. This depends on both age and the number of children.

Several studies have found that age of the women at the first birth is important for the risk of mainly breast cancer. The age of the first birth is highly correlated with the number of children, Figure 1, particularly when there are many children, making it difficult to include this covariate in the model in addition to a covariate for the number of children. Instead, it is introduced a separate covariate for each number of children of the women:

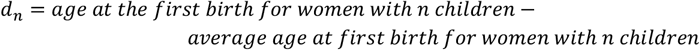

This eliminates the covariation problem. The covariate *d*_*n*_ only has effect on women with n children. This makes the covariates number of children, n, and the value of *d*_*n*_ when it is non-zero, independent of each other. The covariates *a, c, d*_1_, *d*_2_, *d*_3_, … . are used in the model:

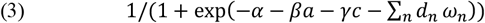

with the parameters *α, β, γ, ω*_1_, *ω*_2_, *ω*_3_, … The data set is so large that the new parameters *ω*_*n*_ can be estimated with small uncertainty for n<8.

First, an analysis was performed with all parameters in each of Model 1-3 corresponding to equations (1-3). Then the parameters that are not significant were removed and another analysis was performed. The parameters were chosen that minimized the Akaike information criterion, AIC=2K-2ln(L). Here K is number of model parameters and L is the likelihood of the model. Minimizing AIC is considered as the best trade-off between few parameters and a poor fit to data and many parameters and a good fit to the variability in the data. It is not possible to compare AIC between different datasets. A decrease of AIC with 2 is considered as a much better fit to the data (19). The p-value for each covariate in the model are reported. Lastly, the reduction in deviance between data and model is shown.

## Results

Table 3 shows the average age when dying by age at in 1960, the start of follow-up period and parity. The mortality seems independent of number of children. Each additional pregnancy does not change the total mortality rates.

The number of cancer diagnoses before age 90 as a function of number of children in the present marriage and the age at the time of diagnosis for the four different cancer diagnosis are given in Tables 5, 6, 7, 8, Upper panel. The number of ovarian and endometrial cancers are almost identical. In the lower panel of the tables are given the number per 10 000 women years (ie. number of diagnoses divided by number of years with women in the category) of the four types of cancer for each age group and number of children.

**Table 5.** Upper panel. Number of **breast cancer** diagnoses occurring in the age range in the 1960 census as a function of number of children in the present marriage. Lower panel. Age-specific incidence rates (per 10.000 PY) of breast cancer after number of children in the present marriage. Rates based on <3 cases not shown.

| Age\chi<br>N.cancers | 0 | 1 | 2 | 3 | 4 | 5 | 6 | 7 | 8 | 9 | 10 | 11 | 12 | 13 | 14 | 15 |
| --- | --- | --- | --- | --- | --- | --- | --- | --- | --- | --- | --- | --- | --- | --- | --- | --- |
| 45-59 | 415 | 560 | 660 | 395 | 214 | 83 | 39 | 22 | 2 | 1 | 2 | 0 | 1 | 0 | 0 | 1 |
| 60-74 | 1364 | 1502 | 1969 | 1170 | 619 | 300 | 157 | 67 | 45 | 22 | 11 | 7 | 5 | 1 | 0 | 0 |
| 75-89 | 1331 | 1461 | 1794 | 1231 | 679 | 333 | 201 | 91 | 72 | 37 | 24 | 7 | 5 | 3 | 1 | 1 |
| Sum | 3110 | 3523 | 4423 | 2796 | 1512 | 716 | 397 | 180 | 119 | 60 | 37 | 14 | 11 | 4 | 1 | 2 |
| Incidence<br>rates by<br>Age | 0 | 1 | 2 | 3 | 4 | 5 | 6 | 7 | 8 | 9 | 10 | 11 | 12 | 13 |  |  |
| 45-59 | 12.7 | 13.7 | 11.8 | 10.7 | 10.3 | 7.9 | 7.3 | 7.8 |  |  |  |  |  |  |  |  |
| 60-74 | 18.6 | 18.9 | 19.0 | 16.4 | 14.3 | 12.4 | 11.3 | 8.2 | 9.0 | 7.4 | 6.0 | 7.7 | 10.0 |  |  |  |
| 75-89 | 27.9 | 28.8 | 26.2 | 25.0 | 21.9 | 18.2 | 18.0 | 12.3 | 15.9 | 13.1 | 13.4 | 7.6 | 9.2 | 13.8 |  |  |
| Sum | 20.3 | 20.6 | 19.4 | 17.8 | 15.9 | 13.5 | 13.1 | 9.9 | 10.8 | 9.1 | 9.0 | 6.8 | 9.5 | 7.9 |  |  |

**Table 6.** Upper panel: Number of **ovarian cancer** diagnoses occurring in the age range as a function of number of children in the present marriage. Lower panel: Age-specific incidence rates (per 10.000 PY) of ovarian cancer after number of children in the present marriage. Rates based on <3 cases not shown.

| Age\chi<br>N.cancers | 0 | 1 | 2 | 3 | 4 | 5 | 6 | 7 | 8 | 9 | 10 | 11 | 12 | 13 |
| --- | --- | --- | --- | --- | --- | --- | --- | --- | --- | --- | --- | --- | --- | --- |
| 45-59 | 149 | 187 | 175 | 95 | 53 | 20 | 11 | 4 | 4 | 1 | 1 | 0 | 0 | 0 |
| 60-74 | 365 | 398 | 471 | 263 | 160 | 66 | 28 | 12 | 7 | 2 | 3 | 2 | 0 | 1 |
| 75-89 | 253 | 232 | 365 | 248 | 110 | 70 | 38 | 18 | 8 | 2 | 3 | 2 | 0 | 0 |
| Sum | 767 | 817 | 1011 | 606 | 323 | 156 | 77 | 34 | 19 | 5 | 7 | 4 | 0 | 1 |
| Incidence<br>rates by<br>age | 0 | 1 | 2 | 3 | 4 | 5 | 6 | 7 | 8 | 9 | 10 | 11 |  |  |
| 45-59 | 4.6 | 4.6 | 3.1 | 2.6 | 2.6 | 1.9 | 2.1 | 1.4 | 2.6 |  |  |  |  |  |
| 60-74 | 5.0 | 5.0 | 4.5 | 3.7 | 3.7 | 2.7 | 2.0 | 1.5 | 1.4 |  | 1.6 |  |  |  |
| 75-89 | 5.3 | 4.6 | 5.3 | 5.0 | 3.5 | 3.8 | 3.4 | 2.5 | 1.8 |  | 1.7 |  |  |  |
| Sum | 5.0 | 4.8 | 4.4 | 3.8 | 3.4 | 2.9 | 2.5 | 1.9 | 1.7 | 0.76 | 1.7 | 2.0 |  |  |

**Table 7.** Upper panel. Number of **endometrial cancer** diagnoses occurring in the age range as a function of number of children in the present marriage. Lower panel; Age-specific incidence rates (per 10.000 PY) of endometrial cancer after number of children in the present marriage. Rates with <3 cases not shown.

| Age\chi<br>N.cancers | 0 | 1 | 2 | 3 | 4 | 5 | 6 | 7 | 8 | 9 | 10 | 11 | 12 | 13 |
| --- | --- | --- | --- | --- | --- | --- | --- | --- | --- | --- | --- | --- | --- | --- |
| 45-59 | 157 | 164 | 182 | 112 | 43 | 19 | 11 | 7 | 3 | 1 | 1 | 2 | 1 | 0 |
| 60-74 | 412 | 399 | 436 | 273 | 152 | 90 | 39 | 16 | 7 | 6 | 0 | 1 | 1 | 0 |
| 75-89 | 258 | 255 | 333 | 196 | 116 | 61 | 32 | 20 | 11 | 7 | 7 | 0 | 2 | 1 |
| Sum | 827 | 818 | 951 | 581 | 311 | 170 | 82 | 43 | 21 | 14 | 8 | 3 | 4 | 1 |
| Incidence<br>rates by<br>age | 0 | 1 | 2 | 3 | 4 | 5 | 6 | 7 | 8 | 9 | 10 | 11 | 12 |  |
| 45-59 | 4.8 | 4.0 | 3.2 | 3.0 | 2.1 | 1.8 | 2.1 | 2.5 | 2.0 |  |  |  |  |  |
| 60-74 | 5.6 | 5.0 | 4.2 | 3.8 | 3.5 | 3.7 | 2.8 | 1.9 | 1.4 | 2.0 |  |  |  |  |
| 75-89 | 5.4 | 5.0 | 4.9 | 4.0 | 3.7 | 3.3 | 2.9 | 2.8 | 2.4 | 2.5 | 3.9 |  |  |  |
| Sum | 5.4 | 4.8 | 4.2 | 3.7 | 3.3 | 3.2 | 2.7 | 2.4 | 1.9 | 2.1 | 2.0 | 1.5 | 3.4 |  |

**Table 8.** Upper panel. Number of cervical cancer diagnoses occurring in the age range as a function of number of children in the present marriage. Lower panel. Age-specific incidence rates (per 10.000 PY) of cervical cancer after number of children in the present marriage.

| Age\chi<br>N.cancers | 0 | 1 | 2 | 3 | 4 | 5 | 6 | 7 | 8 | 9 | 10 | 11 | 12 | 13 |
| --- | --- | --- | --- | --- | --- | --- | --- | --- | --- | --- | --- | --- | --- | --- |
| 45-59 | 153 | 150 | 211 | 134 | 80 | 43 | 21 | 14 | 9 | 3 | 1 | 2 | 1 | 1 |
| 60-74 | 262 | 276 | 304 | 176 | 119 | 63 | 35 | 29 | 17 | 14 | 5 | 3 | 2 | 4 |
| 75-89 | 108 | 95 | 132 | 88 | 52 | 38 | 23 | 14 | 9 | 11 | 0 | 3 | 2 | 0 |
| Sum | 523 | 521 | 647 | 398 | 251 | 144 | 79 | 57 | 35 | 28 | 6 | 8 | 5 | 5 |
| Incidence<br>rates by<br>age | 0 | 1 | 2 | 3 | 4 | 5 | 6 | 7 | 8 | 9 | 10 | 11 | 12 | 13 |
| 45-59 | 4.7 | 3.7 | 3.8 | 3.6 | 3.9 | 4.1 | 3.9 | 4.9 | 5.9 | 3.7 |  |  |  |  |
| 60-74 | 3.6 | 3.5 | 2.9 | 2.5 | 2.8 | 2.6 | 2.5 | 3.5 | 3.4 | 4.7 | 2.7 | 3.3 |  |  |
| 75-89 | 2.3 | 1.9 | 1.9 | 1.8 | 1.7 | 2.1 | 2.1 | 2.0 | 2.0 | 3.9 |  | 3.3 |  |  |
| Sum | 3.4 | 3.0 | 2.8 | 2.5 | 2.6 | 2.7 | 2.6 | 3.1 | 3.2 | 4.2 | 1.5 | 3.9 | 4.3 | 9.8 |

The reduction in incidence rates for cancer with increasing number of children 0 till 14 is illustrated in Figure 2 with a (0.05,0.95) confidence interval for the probability per year for the four cancer sites. The same linear decrease is observed for all three HDC cancers. There is no reduction with increasing number of children for cervical cancer. There is larger uncertainty and some noise in the curves for higher number of children due to less data. However, for the sum of the three HDC cancers the uncertainty is small up to 14 children as shown in Figure 3. Figure 2 shows that the ratio of cancer diagnoses for women without children is between the ratio for women with one and two children for breast cancer but not for the two other cancers.

**Figure 2.**
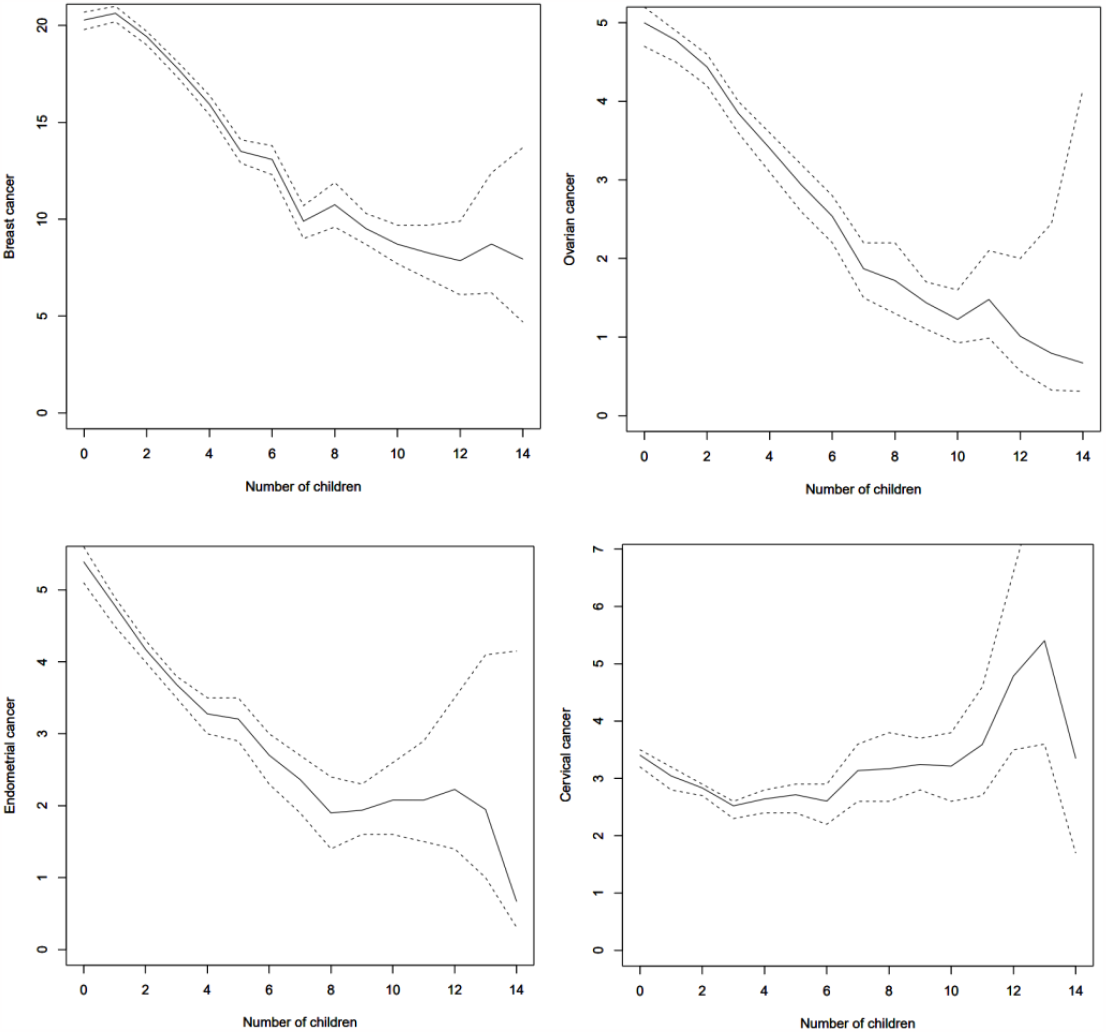
The incidence rate per 10 000 person-years for breast, ovarian, endometrial, and cervical cancer as a function of number of children 0-14. The dotted lines are the estimated (0.05,0.95) confidence interval.

**Figure 3.**
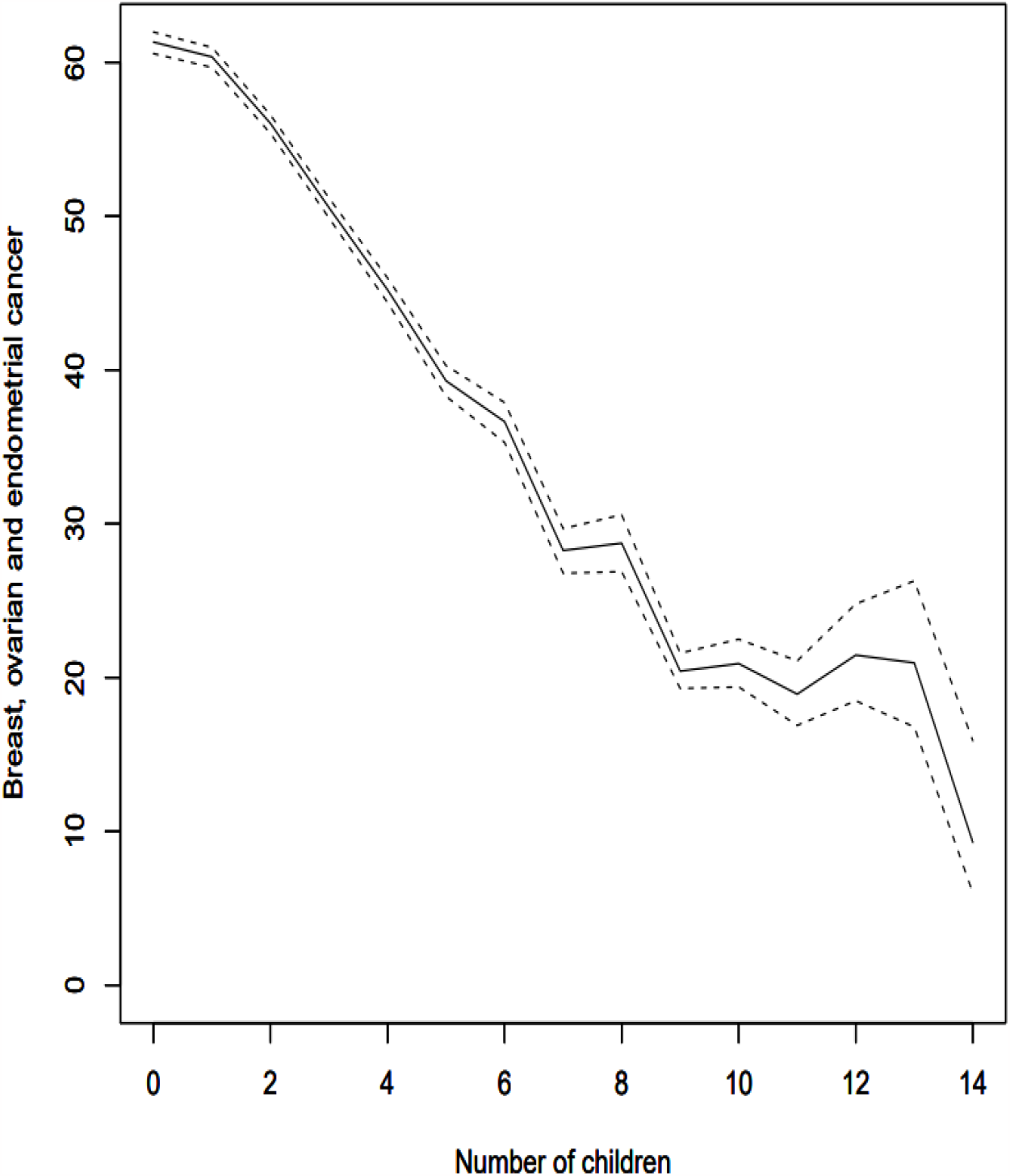
The incidence rate per 10 000 person-years for the sum of breast, ovarian and endometrial cancer as a function of number of children 0-14. The dotted lines are the estimated (0.05,0.95) confidence interval.

### Logit regression analysis

The probability for the four cancers was modelled using a logit regression model with age and number of children as covariates. Women without children were omitted. The results for the three Model 1-3 and the four types of cancer and the sum of the three HDC are shown in Table 9-13 and Figure 4-8. For breast, ovarian and endometrial cancer, and the sum of the three HDC, a good fit with the linear model (1) was obtained for the probability of cancer as a function of age and number of children. All parameters are significant with p<1 e-11.

**Table 9.** The estimated parameters for breast cancer in Model 1-3. Linear model: estimated AIC equal 200783, deviance reduced from 202392 to 200777. Higher order model: estimated AIC equal 200777, deviance reduced from 202392 to 200767. Corresponding numbers are not comparable in the model with age at birth since the data set is smaller.

| Param. | Linear model, Model 1 |  |  | Higher order model, Model 2 |  |  | Age at first birth, Model 3 |  |  |
| --- | --- | --- | --- | --- | --- | --- | --- | --- | --- |
|  | Est. | S.dev. | P | Est. | S.dev. | P | Est. | S.dev. | P |
| $\alpha$ (interc.) | -8.013 | 0.060 | <2e-16 | -8.663 | 0.37 | <2e-16 | -8.01 | 0.061 | <2e-16 |
| $\gamma$ (child) | -0.111 | 0.0052 | <2e-16 | -0.197 | 0.039 | 5e-7 | -0.111 | 0.0052 | <2e-16 |
| $\beta$ (age) | 0.0283 | 0.00083 | <2e-16 | 0.0508 | 0.011 | 1e-6 | 0.028 | 0.00083 | <2e-16 |
| $\theta$ (age**2) | | | | -0.00018 | 0.000075 | 0.015 | | | |
| $\varphi$<br>(age*child) | | | | 0.0012 | 0.00053 | 0.026 | | | |
| $\omega_1$ | | | | | | | 0.011 | 0.0021 | <2e-7 |
| $\omega_2$ | | | | | | | 0.0095 | 0.0043 | 0.03 |

**Figure 4.**
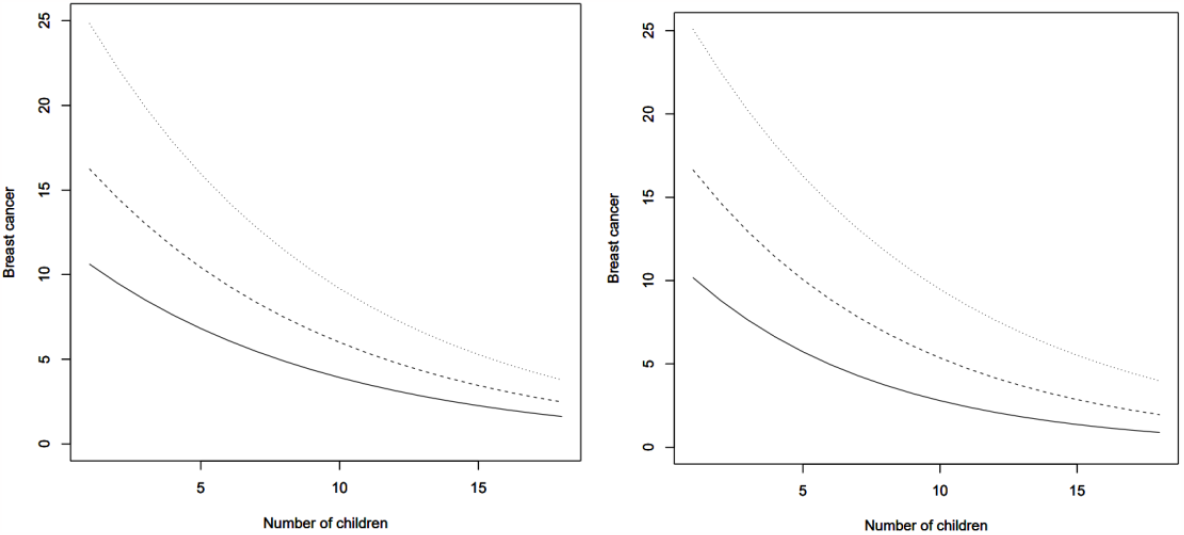
The incidence rate per 10 000 women for **breast cancer** at the age 45 (lower curve), 60 and 75 (upper curve) as a function of number of children. Left the linear model, right including higher order terms.

**Figure 5.**
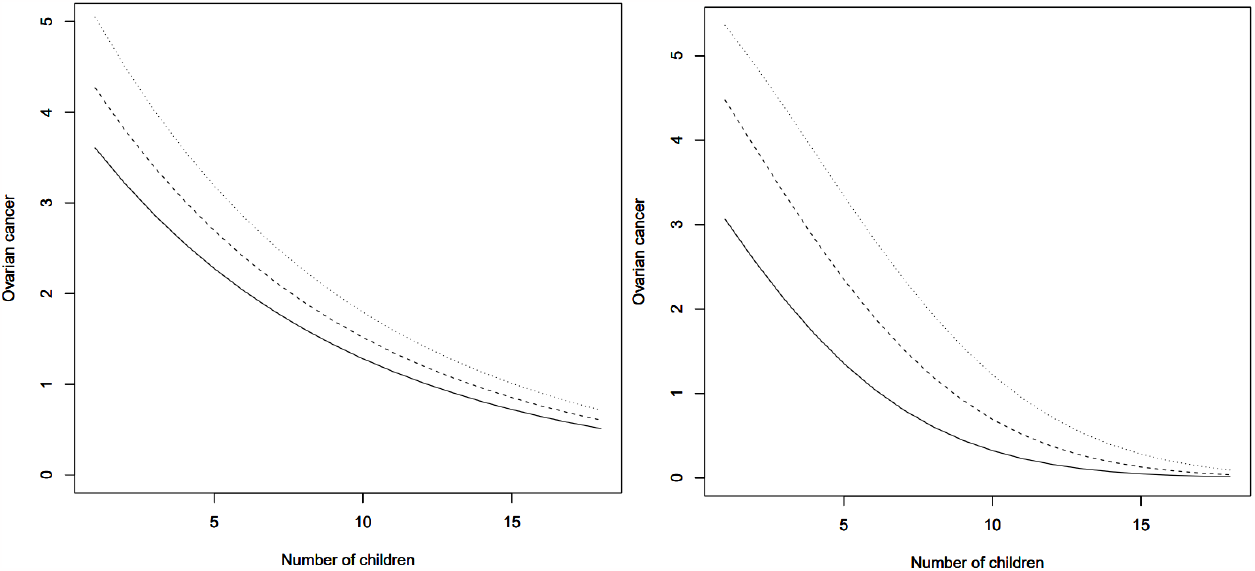
The incidence rate per 10 000 women for **ovarian cancer** at the age 45 (lower curve), 60 and 75 (upper curve) as a function of number of children in present marriage at the 1960 census. Left the linear model, right including higher order terms.

**Figure 6.**
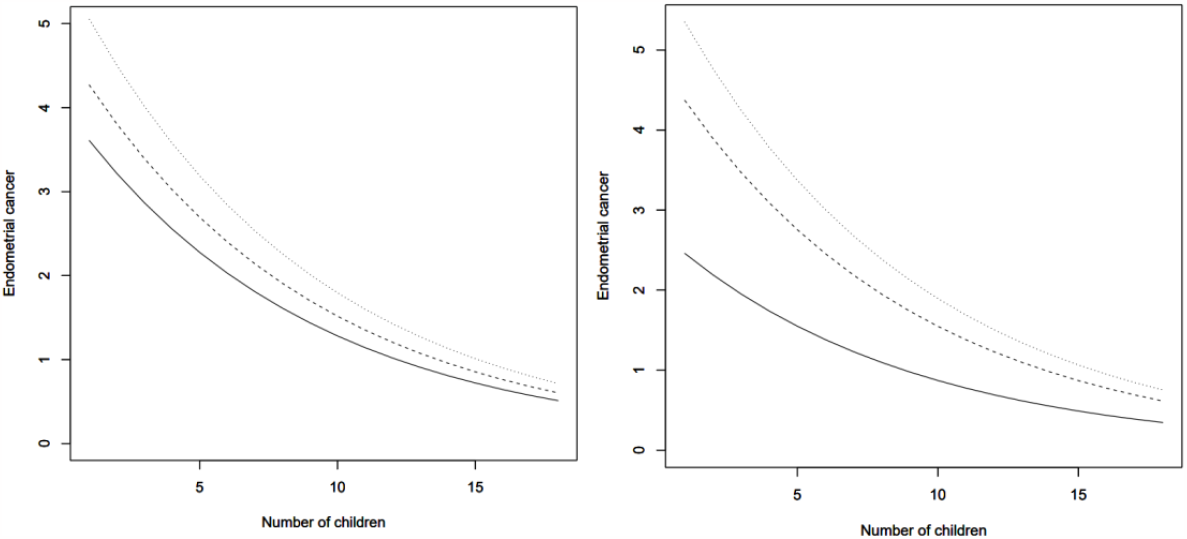
The estimated incidence rate per 10 000 women for **endometrial cancer** at the age 45 (lower curve), 60 and 75 (upper curve) as a function of number of children in present marriage at the 1960 census. Left figure a linear model, right including higher order terms.

**Figure 7.**
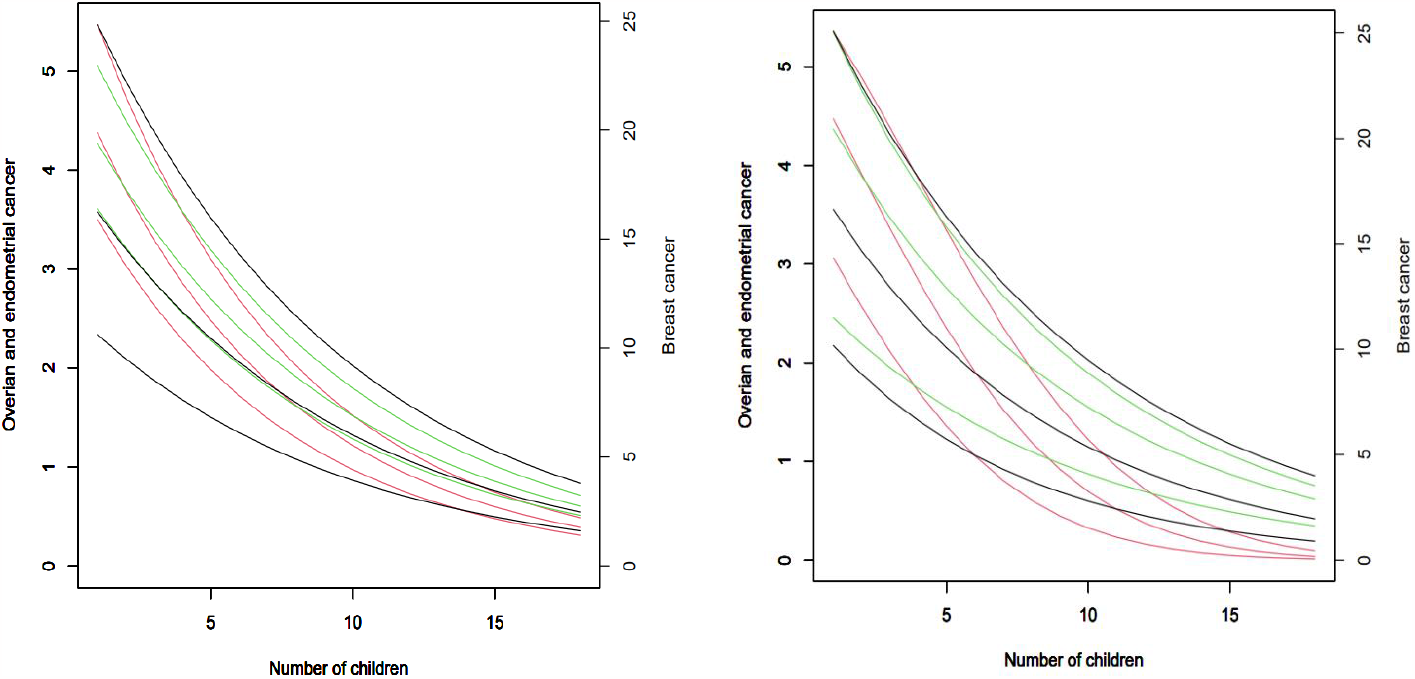
The estimated incidence rate per 10 000 women for cancer of the breast (black, right axis), ovary (red, left axis) and endometrium (green, left axis) for the ages 45, 60 and 75 years for 1 until 18 children. Left figure linear model right figure higher order terms.

**Figure 8.**
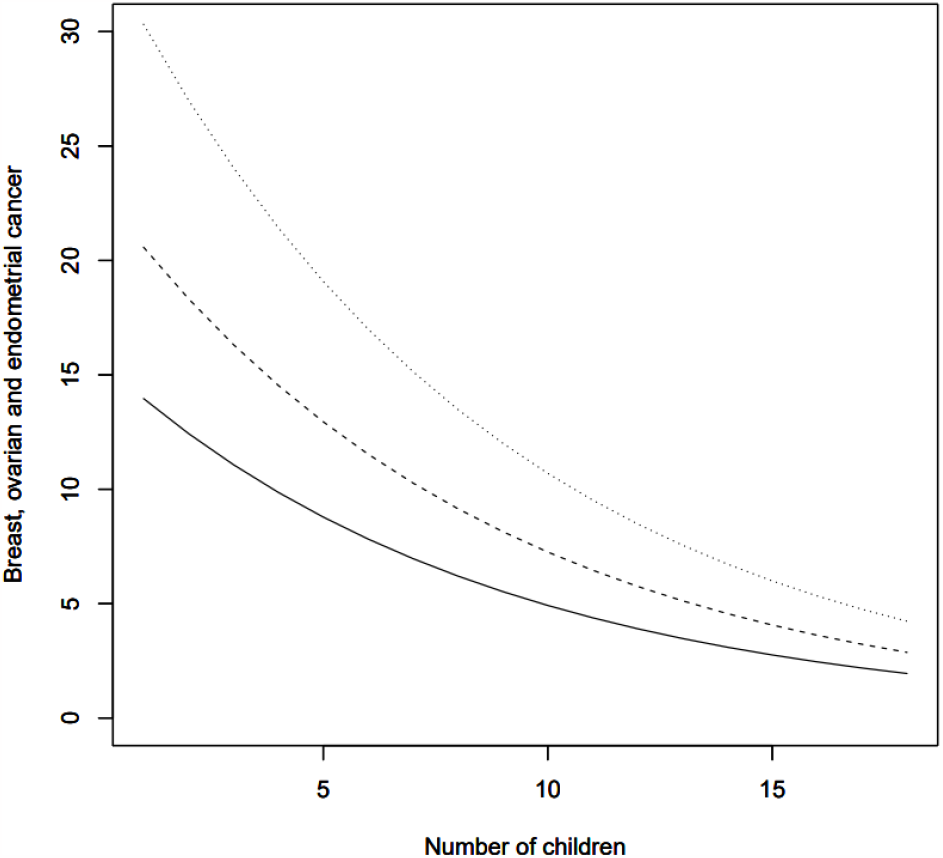
The ratio per 10 000 women for the sum of **breast, ovarian and endometrial cancer** at the age 45 (lower curve), 60 and 75 (upper curve) as a function of number of children in present marriage at the 1960 census. Linear model. The higher order model did not give a better fit to the data and is therefore not included.

The probability for cancer is reduced for each additional child. The reduction in relative probability, given the estimated parameter 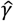, is 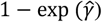, or 10.5% (95% CI; 9.6-11.4) per child for breast cancer, 13.2% (11.2-15.3) for ovarian cancer, and 10.9% (8.9-12.8) for endometrial cancer based on the estimated parameters and their uncertainty.

This implies that the relative probability for breast cancer with n+1 children is 10.5% less than for n children for n>0. In this model, this decrease is independent of number of children. This gives the risk reduction (RR) for 2 children compared to 1 child equal to 0.90 (95% CI 0.89, 0.90), 0.87 (0.85, 0.89), 0.89 (0.87, 0.91) and 0.89 (0.88, 0.90) respectively for the three HDC cancers and the sum of the three cancers, for 5 children compared to 1 child 0.64 (0.62, 0.67), 0.57 (0.52, 0.61), 0.63 (0.58, 0.69) and 0.63 (0.61, 0.65) respectively, for 10 children compared to 1 child 0.37 (0.34, 0.40), 0.28 (0.23, 0.34), 0.36 (0.29, 0.43) and 0.35 (0.32, 0.38) respectively, and 0.24 (0.21, 0.27), 0.16 (0.12, 0. 21), 0.22 (0.17, 0.30) and 0.22 (0.20, 0.25) respectively for 14 children compared to 1 child. These confidence intervals are based on the linear logit model using data from all women with children and gives slightly different values than the intervals illustrated in Figure 2 and 3 where we use a binominal model and the estimate for n children is based on the data for only for n children (n low) or data for n-1, n, n+1 and n+2 children (n high). The binominal model gives larger CI-intervals for high parity since the data is sparse.

In a model with higher order terms, Model 2, all terms were included at first. Then the terms with p>0.05 were removed and a new analysis performed. Notice that the added terms have small coefficients and do not change the main picture. Model 2 reduces the AIC and is therefore considered as a better model for the variability in the data, but the uncertainty in the parameter estimation increases compared to the result from Model 1. The curvilinear results are presented for the HDC in Figures 4, 5, 6, 7. The left panels show estimates from Model 1, the right panel from Model 2. The curves are quite similar the results with only linear terms. The main difference is that the curve for age equal 60 years is lower for ovarian and endometrial cancer. With higher order terms added the reduction in probability for each additional child varies both with number of children and age. In Model 2 is the reduction dependent of both age and number of children. The reduction in probability for breast cancer per added child in the preferred model is 10% at age 60 and increases to 13% at age 75 and is independent of number of children. For ovarian cancer the reduction in the preferred model is when increasing from one to two children 8.6% at age 60 and 16% at age 75 and when increasing from 5 to 6 children the reduction is 15% and 22% respectively. For endometrial cancer the reduction in probability for cancer per added child is 10.9% independent of age and number of children.

For the sum of the three HDC cancer, the parameter estimates are shown in Table 12 and the curves in Figure 8. The general properties of the three HDC cancers are very similar except that breast cancer has about 4.4 times as high probability. When including higher order terms, Model 2, the result with the three HDC cancers is quite like the results with only breast cancer. Two higher order term have p<0.05 as reported in Table 12. The higher order model did not give a reduction in AIC nor deviance and is therefore not included in Figure 8.

**Table 10.** The estimated parameters for ovarian cancer in Model 1-3. Linear model: AIC estimated equal 53886, deviance reduced from 54115 to 53880. Higher order model: estimated AIC equal 53876, deviance reduced from 54115 to 53864. Corresponding numbers are not comparable in the model with age at birth since the data set is smaller.

| Param. | Linear model, Model 1 |  |  | Higher order model, Model 2 |  |  | Age at first birth, Model 3 |  |  |
| --- | --- | --- | --- | --- | --- | --- | --- | --- | --- |
|  | Est. | S.dev. | P | Est. | S.dev. | P | Est. | S.dev. | P |
| $\alpha$ (interc.) | -8.485 | 0.123 | <2e-16 | -10.13 | 0.76 | <2e-16 | -8.534 | 0.124 | <2e-16 |
| $\gamma$ (child) | -0.142 | 0.012 | <2e-16 | -0.277 | 0.089 | 0.0018 | -0.138 | 0.0012 | <2e-16 |
| $\beta$ (age) | 0.0149 | 0.0017 | <2e-16 | 0.0687 | 0.0022 | 0.00151 | 0.0151 | 0.00089 | <2e-16 |
| $\mu$ (child**2) | | | | -0.00922 | 0.00045 | 0.039 | | | |
| $\theta$ (age**2) | | | | -0.000441 | 0.00016 | 0.0055 | | | |
| $\varphi$<br>(age*child) | | | | 0.00286 | 0.00122 | 0.020 | | | |
| $\omega_1$ | | | | | | | -0.0334 | 0.0046 | 6e-13 |
| $\omega_2$ | | | | | | | -0.0479 | 0.0098 | 9e-7 |

**Table 11.** The estimated parameters for endometrial cancer in Model 1-3. Linear model: estimated AIC equal 53139, deviance reduced from 53283 to 53133. Higher order model: estimated AIC equal 53114, deviance reduced from 53283 to 53106. Corresponding numbers are not comparable in the model with age at birth since the data set is smaller.

| Param. | Linear model, Model 1 |  |  | Higher order model, Model 2 |  |  | Age at first birth, Model 3 |  |  |
| --- | --- | --- | --- | --- | --- | --- | --- | --- | --- |
|  | Est. | S.dev. | p | Est. | S.dev. | P | Est. | S.dev. | P |
| $\alpha$ (interc.) | -8.317 | 0.124 | <2e-16 | -12.16 | 0.77 | <2e-16 | -8.321 | 0.124 | <2e-16 |
| $\gamma$ (child) | -0.115 | 0.011 | <2e-16 | -0.115 | 0.011 | <2e-16 | -0.114 | 0.0011 | <2e-16 |
| $\beta$ (age) | 0.0112 | 0.0017 | 1e-11 | 0.0126 | 0.023 | 3e-8 | 0.0112 | 0.0017 | 1e-10 |
| $\theta$ (age**2) | | | | -0.000830 | 0.00016 | 4e-7 | | | |
| $\omega_1$ | | | | | | | -0.0104 | 0.0046 | 0.023 |

**Table 12.** The estimated parameters for the sum of breast, ovarian and endometrial cancer in Model 1-2. Linear model: estimated AIC equal 238722, deviance reduced from 240526 to 238710. Higher order model: estimated AIC equal 238726, deviance reduced from 240526 to 238716. Note that AIC og deviance is higher in linear model. The model is included since two higher order parameters were significant.

| Param. | Linear model, Model 1 |  |  | Higher order model, Model 2 |  |  |
| --- | --- | --- | --- | --- | --- | --- |
|  | Est. | S.dev. | P | Est. | S.dev. | P |
| $\alpha$ (interc.) | -7.622 | 0.054 | <2e-16 | -8.373 | 0.33 | <2e-16 |
| $\gamma$ (child) | -0.116 | 0.0047 | <2e-16 | -0.222 | 0.035 | 4e-10 |
| $\beta$ (age) | 0.0259 | 0.00074 | <2e-16 | 0.0521 | 0.0095 | 4e-8 |
| $\theta$ (age**2) | | | | -0.00021 | 0.000068 | 0.016 |
| $\varphi$ (age*child) | | | | 0.0015 | 0.00048 | 0.026 |

**Table 13.**
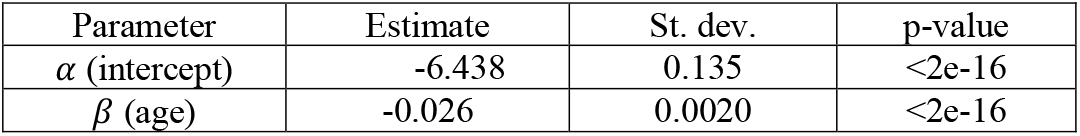
The estimated parameters for cervical cancer in Model 1. AIC estimated to 39936.

In Model 3 all covariates for the age at birth of the first child were included at first and then performed a second modelling leaving out covariates with p>0.05. For breast and ovarian cancer, the covariates for 1 and 2 children were significant and for endometrial cancer only the covariate for age at the first child was significant. The results are presented in Tables 9-11, right panel. For the sum of all three HDC cancers the covariate with age at first birth was not significant and therefore not included in Table 12. Table 2 shows that 80% of the women with one child have an age in the interval (22,37) years with 28 years as average. The corresponding intervals for two and three children are (21,34) and (21,32). The birth 5 years earlier with a covariate equal 0.014 implies a reduced risk for cancer equal exp (-5*0.014) =0.93. This is only 70% of the reduction due to one more child which is 0.9.

The corresponding analysis for cervical cancer, Table 13, shows that the probability for cervical cancer decreases with age and is independent of number of children. The regression term for children was far from significant. For cervical cancer the curves are horizonal lines since the probability is independent of parity (not shown).

## Discussion

We found highly significant and similar curvilinear relationships between number of full-term pregnancies and each of the three hormone-dependent cancers over the full fertility range up to 18 children. The figures clearly demonstrate the dominating relationship of parity for the three cancer sites separately and combined.

In a simple linear, logit model, the reduction in incidence rates was around ten percent for each child for breast and endometrial cancer and slightly higher for ovarian cancer. Second order terms increased the fit for the model but reduced the precision of the estimates. The higher order model gave a stronger effect of parity on ovarian cancer while for breast cancer age became more important. No changes were shown for endometrial cancer. The figures illustrate how the seemingly small changes of around ten percent reduction for each child cumulate to an impressive effect throughout life for grand parity women.

No effect was found for cancer of the cervix which is known to have other risk factors like HPV virus (18). The total mortality was found to be the same regardless of parity. This is compatible with previous work (10,12) where the reduced number of deaths from hormone-dependent cancers in high parous women are balanced against more deaths of other diseases.

The new statistical approach for the analysis of the collinearity of parity and age at first birth showed that an early age at first birth, here with age at marriage as proxy, had less effect than another child on the incidence rates of all three cancer sites. The joint model with parity and age at first birth as almost independent variables shows the combined effect of the two covariates. This is to our best knowledge a novel approach.

The interpretation of the findings must consider the similarity of the shapes of the curves for three different cancer sites and the strong protective effect in grand-grand multiparous postmenopausal women. The strong linear relationships make it almost impossible to explain the findings by any confounder. This was thoroughly discussed for the first results linking cigarette smoking to lung cancer and the relation to other proposed risk factors at that time (20). In an updated summary of the causality criteria both temporality and dose-response are still the core of causality criteria (21). The findings could be explained by two different hypotheses:

The first hypothesis would focus on the separate causes for each of the three HDC. This hypothesis would imply different major, strong risk factors for each of the three cancer sites. No risk factors or exposures are proposed that could exhibit such confounding risks in each of the three HDC sites. One could argue that for each site there could be different risk factors. This would demand a very complicated set of biological theories.

The second hypothesis propose that the incidence rates of the three HDC are related to full-term pregnancies as the major cause for all three sites.

It is important to note that the protective effect of the pregnancy is linked to full-term pregnancies 34 weeks or more (22). This explanation for the effect of high parity on breast cancer incidence has been put forward as the semi-allograft theory (23). This theory relies on the well-known dynamic changes during pregnancies of the immune systems of fetus and mother (24, 25, 26). The hypothesis is based on the findings that each full-term pregnancy has the same effect and not only the first as proposed by others (27). The future research could focus on the immunological mechanisms behind these effects. Abortions have not been found to protect against breast cancer (28, 29).

The discussion of potential confounders in this study must take into considerations that these birth cohorts born 1871 till 1915 had highly different lives from today’s Norwegian women. Norway was a very poor country with a large emigration to the US. Poverty was extensive both in cities and on the countryside leaving a population with a low average BMI. The abuse of alcohol among men was extreme. This gave a full stop of legal hard liquor during the first world war and for ten years after. Women had a low consumption of alcohol. Smoking was not at all common among women even up to 1960. The study cohort had no access to OC or HRT. Women were expected to be married before the first birth and assumed to stay at home while the husband was working. Women with a child outside marriage was looked upon by many as a shameful woman (30). Consequently, a substantial part of the female population stayed throughout life as nulliparous. The fertility rates for the different birth cohorts included here, 1871 to 1915, differed over time and so did age at first birth.

### Some methodological moments

#### Use of observed versus expected values from the population

Several of the studies of high or grand parity have compared the observed number (O) of cancers with expected (E) number based on the use of national incidence or mortality figures (6. 7, 10, 11). The expected value is an estimate of the average risk of a specific cancer site in the population given the existing average parity. Average number of children in the different populations analyzed could be between 2 and 3 children. The observed versus expected calculations O/E have given reduced or flawed risk estimates for multiparous women due to the undefined reference group. However, in smaller studies will the use of national expected values add statistical power to the estimation of risk due to the expected values from the total population.

### Collinearity

Studies of hormone-dependent cancers and parity mostly includes different aspects of parity like number of children, age at first and last birth, and lactation. Little attention has been paid to the fact that these variables are dependent on each other. The quantile of age at present marriage registered in the 1960 census for a given number of children is given in Table 2.

When parity increases, the range of age at first and last birth diminish as shown in Figure 1. The findings in this analysis of a continuous downward trend in the incidence rates for ten or more children clearly demonstrates the independent effect of high parity on the incidence rates for all three cancer sites. The use of multiplicative models like Cox analyses that implicitly assumes independence between covariates, could be misleading. If there are no observed values for certain combinations of the exposure variables an expected value will be created anyhow. This could have important effects on estimations of marginal values in the observed data. As an example, age at first birth has a restricted range for multiparous women. There are almost no observations of age at first birth >30 if parity is ten or more, see Table 2. Therefore, the logit Model 3 is introduced that decouples the two covariates, parity, and age at first birth.

#### Right-truncation of high parous women

In a dataset with a limited number of women with high parity, it is tempting to treat all high parity women in one group when analysing whether the age of the first birth influence the risk of cancer. This is illustrated in Table 14. Women are divided into groups with women with 1, 2, 3 and >3 children. There is an increasing p-value for 1, 2 to 3 children (2e-7, 0.3, 0.15 respectively) but for the group with >3 children, the p-value is small (0.0015). The reason for the small p-value is that there is a strong correlation between parity and age at first birth. Hence, the association of lower risk for lower age at first birth is caused by the higher parity, not because the early first birth. In a dataset with many women with 4-14 children there is no need to group the high parous women in one group to get significant statistical strength.

**Table 14.** The estimated parameters for breast cancer in Model 3 when women with 4 and more children are collected in one group 4+.

| Param. | Age at first birth |  |  |
| --- | --- | --- | --- |
|  | Est. | S.dev. | P |
| $\alpha$ (interc.) | -8.00 | 0.061 | <2e-16 |
| $\gamma$ (child) | -0.111 | 0.0052 | <2e-16 |
| $\beta$ (age) | 0.028 | 0.00083 | <2e-16 |
| $\theta$ (age**2) | | | |
| $\varphi$ (age*child) | | | |
| $\omega_1$ | 0.011 | 0.0021 | <2e-7 |
| $\omega_2$ | 0.0095 | 0.0044 | 0.03 |
| $\omega_3$ | 0.0091 | 0.0064 | 0.15 |
| $\omega_{4+}$ | 0.021 | 0.0068 | 0.0015 |

Most previous studies have used a collapsed last group i.e., 12-20 children as an example. While the middle value is 16, the arithmetic mean is slightly below 13 which can be calculated based on Table 1. This might give a false impression of the incidence curves flattening out in high parous women. The same applies to the use of right truncations like 5+ or 8+. In the present analyses this is accounted for, see Statistical methods. This correction of the estimated number of women in each parity group will move the points towards left and give a steeper downward curve demonstrating more precisely the effect of each single pregnancy in high parity women.

### Previous studies comparing the three hormone-dependent cancers within the same design

Only a few other studies have published incidence analyses comparable to the present. In a Norwegian prospective study based on population samples from counties in Norway (6) with 63 000 women, 5+ versus 1 child had a relative risk for breast cancer of 0.54, ovary cancer 0.43 and endometrial cancer 0.42 based on the O/E method. Based on Finnish registry data (7) risk estimated for grand-grand multiparity women used standard incidence ratios (SIRs). Among 86 978 GGM women 92 breast cancer, 15 ovarian cancers and 14 endometrial cancers were identified. The respectively SIRs were 0.44, 0,49 and 0.31. Grand multiparity, 5+ children, was investigated in the Jerusalem Perinatal Study Cohort using Cox proportional hazard (8). Participants were 8 246 GM women and 19 703 women with parity 1-4 as reference. The adjusted hazard ratio was 0.62 for breast cancer, for ovarian 0.83, and for endometrial 0.75.

Mortality studies started with a study based on English death certificate questions on parity for 1.2 million deaths between 1938 and 1960, women aged 45-74 years (5). The analysis looked only at nulliparous versus parous women. Mortality ratios were calculated for three different periods. The last period 1959-60 showed age-standardized mortality ratios (SMRs) of 92 for breast cancer, 73 for ovary and 74 for endometrial cancer. In 1990 a Norwegian linkage study between the 1970 Census and the register of deaths in the Central Bureau of Statistics in Norway (9) were undertaken with a total of 6 979 cancer deaths of HDC. In an analysis combining the three cancer sites into one as outcome the mortality decreased linearly with increasing number of children. RR for women with 12-20 children was 0.23 with only 2 deaths. The linear trend gave a reduction in the relative risk for each child of 9.6% (8.3-10.9). No consistent trend with age at first birth were found. In separate analyses for each cancer site the percentage reduction was 7.2% for breast, 12.7% for ovary and 12.2% for endometrial cancer. In 2005, a Finnish study based on parity information in the national population register women found 87 922 women of whom 3 678 had ten or more deliveries (10). Linkage to the Finnish national cause-of-death files gave follow-up out 2001. Standard mortality ratios, SMRs, calculated for all causes of cancer showed that grand-grand multiparous women had SMR for breast cancer of 0.70, ovarian cancer 0.43 and endometrial 0.34 with 24, 6 and 2 deaths. In a study from Taiwan all grand-multiparous women registered in the Birth Register between 1978-87 were included, 144 922 women (11). Analyses were based on SMRs or O/E with a total of 394 breast, 72 ovarian and 18 endometrial cancers. The SMRs for among GM women were 0.73 for breast cancer, 0.64 for ovary and 0.54 for endometrial cancer. Lastly, the census-based Israel Longitudinal Mortality Study II identified 1995-2004 a total of 68 822 women 45-89 years old (12). Record linkage to death registers identified a total of 568 cases of hormone-dependent cancers. Older women, 65-89 years, with 8+ children had a reduced risk in a Cox analysis of 0.22 for reproductive cancers combined or a linear reduction of 10%.

### Studies of parity in single cancer sites

The number of studies of parity and each of the three cancer sites published separately has been extensive over the decades.

Most studies are found for breast cancer. In a prospective study in Norway with 63 000 women (31) a consistent inverse trend with parity was described. Postmenopausal women with 9-15 children had an O/E ratio of 0.30. A Swedish study (32) used census information and registers. The nested case-control analyses had 12 782 incident cases and a 5:1 matching. RR for 1 versus 9 children was 0.68 based on four incident cases. The linear trend was 10% per child, but the trend analysis showed that women with 8 and 9 children had higher RR than those with lower parity. Another large collaborative study (33) with 47 epidemiological studies including 50 302 breast cancer cases with an average age of 50.1 years, found a decrease of 7.0% per child and an additional decrease of 4.3% for every 12 months of breastfeeding. Less than ten percent of the cases were from cohort studies. In the Nurses’ Health Studies (34) breastfeeding were inversely related to hormone-negative receptor breast cancer. The highest parity group was 4+. In the EPIC cohort (35) women with 3+ children as cutoff point had a reduced risk of 0.77. In the Norwegian Women and Cancer Study, NOWAC, the linear trend from 0-6 children was estimated to 8.0% per child (36). Adjustment for major risk factors including the effect of lactation, did not change the estimates. Women with 6 children had a crude relative risk of 50% compared to 49% after adjustment.

Parity in relation to ovarian cancers have been analyzed in the Million Women Study (37) based on follow-up of 1.1 million UK women with 8 719 ovarian cancers. Relative risk for each birth among parous women were 0.94 from 1 till 4+ women. In an analysis from the Ovarian Cancer Consortium (38) 21 prospective studies with 1.3 million women 5 584 ovarian cancers showed a reduction in risk of all ovarian cancer of 0.90 per one child to 4+. Four children had a reduced risk of 0.58. In a meta-analysis of age at last birth and epithelial ovarian cancer (39) information from 2,451,071 women with 19,959 cases age at last birth were negatively associated, 0.77 with no change when adjusted for parity (36).

In a study of endometrial cancer in the EPIC study (40) the relative risk for one versus 4+ children were 0.65. In an Asian analysis of 13 prospective cohort studies pooled was the hazard ratio for 5 deliveries versus nulliparous was 0,37 (41). In a pooled analysis of pregnancy outcome and risk of endometrial cancer in 11 cohort and 19 case-control studies odds ratio was estimated to 0.85 per full-term pregnancy. For women with 9+ children the OR was 0.27.

Overall, there is quite consistent findings in single cancer studies that increasing parity reduces the risk of breast, endometrial and ovary cancers in many different populations. Common to most published analyses the number of studies with grand-grand multiparous women is scarce. None of the described studies demonstrated the linear relationships between the full range of pregnancies and HDC.

### Weaknesses

The discussion of the findings should consider the potential for confounding. In the 1960 Census of women above 15 years 61.8% were married, 26.0% unmarried, 0.4% separated, 1.7% divorced and 10.1% widower. Women without children may be a mix of different groups including women that are not able to give birth, women married at late age in the present marriage and possibly with children in a previous marriage. Divorces were very rare in this period. Some women remarried after becoming a widow or after a divorce. A separate analysis of the age at time of marriage in the present marriage showed that nulliparous women were much older than the parity women at time of marriage. The nulliparous women were taken out of the analyses.

No information on breast-feeding were collected in the census. The discussions should carefully consider the collinearity problem as shown for the age at first birth here. In a reanalysis of 47 epidemiological study the effect of parity was estimated to 7.0% and an independent effect of breastfeeding of 4.3% for each 12 months (33). The Norwegian Women and Cancer study is a representative population-based cohort with 154 000 participants, adjustment for total months of lactation did not change the estimates for number of children in a multivariate adjusted analysis (36). In an analysis from the AMBER consortium (43) parity had no protective effect on receptor positive (ER+) with OR 0.92 (0.81-1.03). Lactation had a protective effect on estrogen receptor ER-negative cancer, OR= 0.81, with no trend for ever breastfed. In the Nurses’ Health Studies (44) parous women who had ever breastfed, had a relative risk of 0.82 (0.74-0.98) for ER-cases (18% of all cases) versus nulliparous, while ER positive cases had no reduced risk. There was no significant trend of breastfeeding for all cases. These large studies could not demonstrate a common reduction in risk for duration of breastfeeding.

No information on use of hormones at and after menopause existed at the time of the census. In Norway the very first preparation for menopausal estrogen replacement was introduces in 1953. The preparation contained etinyloestradiol (0.05mg). Preparations with a progestin have been available since 1960, but the use was seldom until the end of the eighties’ (less than 30 000 users) (45).

Routine use of receptor information in breast cancer started in Norway around 2000 and no information existed for most of these cases.

### Strength

The strength of the study is the statistical power using a whole nation with a high parous population, information collected by civil servants during the same time, and the introduction of the birth number. The data quality is supposed to be good as it is based on compulsory participation in the census with interview information.

Norway has a well-established cancer registry operating from 1953 giving a follow-up of 45 years. The register is based on compulsory reporting of all diagnoses of cancer from the responsible department by originally a standard form. In addition, the register received independently copies of all histopathological diagnoses from the pathology departments. A third source of information was linkage to the register of death certificate diagnoses. For the years 1959-60, the cancer diagnosis was histologically confirmed for 94.1% in breast, 99.1% in corpus uteri and 89.2% in ovary. In an evaluation in 1976 of the completeness of the cancer registry 97.5% of all diagnoses registered in the national hospital system was found in the cancer registry (Lund 1981). In Norway access to diagnostics and treatment of cancer is free of charge.

Due to the large number of grand-grand parity women the use of right truncation like 8+ or 12-20, was avoided.

The introduction of an analytical approach of collinearity improved the understanding of the relationship between increasing parity and age at first birth. Using the 1960 Census with information on only parity and not age at each birth in studies of breast, endometrial and ovarian cancers can be defended by the decreasing event space with increasing parity. The fertile period for each woman is from 18 till 43 years or 25 years of menstruation. To have ten children and pregnancy and lactation period around 2 years or maybe less, 10 children will take around 20 years of the 25. This will reduce the importance of age at first and last birth due to the strong restrictions of possible values. With 12 children or more the two variables will be almost fixed as illustrated in Table 2.

## Conclusion

The 1960 Census represents generations with high fertility in Norway. This large historical cohort made it possible to estimate the risk from one till 18 children also for ovarian and endometrial cancer due to sufficient statistical power. A new statistical method was used for the combined analysis of the collinearity between parity and age at first birth (marriage).

We have shown for the first time a strong curvilinear reduction in the incidence rates for all three cancer sites separately and combined for each additional pregnancy or child, over the whole exposure range of 1-18. The simplest explanation for the similarities of the three curves is a common cause – the pregnancies. The terms parity or number of children have potentially distracted the biological thinking away from the immunological changes in each pregnancy.

These findings support the hypothesis that reduced fertility is the major, strong etiological factor for the three hormone-dependent cancers in postmenopausal women. Further studies are needed to explore this hypothesis that the common protection is an effect of similar changes in the immune system during each pregnancy.

## Data Availability

Most of the data is presented in the article. The complete data set has limitations due to privacy and only available in an application to Statistics Norway, Cancer Registry of Norway and Norwegian Institute of Public Health

## Abbreviations

HDC: hormone-dependent cancer
SMR: standard mortality ratio
OR: odds ratio
RR: relative risk
GM: grand multiparity
GGM: grand-grand multiparity
PY: person years of follow-up

## Privacy

The restriction on the information was a follow-up till 90 years, no geographical information and all published results based on less than five persons should be truncated.

The women in the Cohort are born in the period 1871-1915. These women are only followed until they are 90 years old witch are at least 18 years ago. All women are dead except for few women in Table 1 and 2 with an age of at least 108 years. Most of the Cohort died at least 30 years ago. There is so little information that it is not possible to identify the women since there is no dates and no geographical information. Only one person (LH) has access to the data. In all published results, there is at least 5 women in each cell.

## Author contributions

EL started the linkage study, initiated this methodological collaboration and raised funding. EL and LH carried out conception and design of the study. EL and LH were involved in the development of methodology. LH performed data analysis (eg, statistical analysis, biostatistics, and computational analysis). EL, L-TRB and LH wrote, reviewed, and revised the manuscript. EL, L-TRB and LH provided administrative, technical, or material support. EL conducted study supervision. LH had the responsibility for data management. All authors contributed toward data analysis, drafting, and critically revising the paper, gave final approval of the version to be published, and agree to be accountable for all aspects of the work.

## Disclaimer

Some of the data in this article are from the Cancer Registry of Norway. The Cancer Registry of Norway is not responsible for the analysis or interpretation of the data presented.

## References

1. Miller AB. An overview of hormone-associated cancers. Cancer Res 1978 Nov;38(11 Pt 2):3985–90. PMID: 359131 Review.

2. B E Henderson ^1^, H S Feigelson. Hormonal carcinogenesis. Carcinogenesis 2000 Mar;21(3):427–33. doi: 10.1093/carcin/21.3.427.

3. Bogen E. The Cause of Breast Cancer. Am J Public Health Nations Health. 1935 Mar;25(3):245–50. doi: 10.2105/ajph.25.3.245.

4. The Lancet. The Search for Causes of Breast Cancer. 1965;286(7415):725–726. doi:10.1016/S0140-6736(65)90461-7

5. Valerie B. Long term effects of childbearing on health. Epidemiol Community Health. 1985 Dec;39(4):343–6. doi: 10.1136/jech.39.4.343.

6. Kvåle G, Heuch I, Nilssen S. Reproductive factors and cancers of the breast and genital organs – are the different cancer sites similarly affected? Cancer Detection Prevention 1991; 15: 369–77.

7. Högnäs E, Kauppila A, Pukkala E, Tapanainen JS. Cancer risk in women with 10 or more deliveries. Obstet Gynecol. 2014 Apr;123(4):811–6. doi: 10.1097/AOG.0000000000000182.

8. Paltiel O, Tajuddin SM, Polanker Y, Yazdgerdi S, Manor O, Friedlander Y, Harlap S, Calderon-Margalit R. Grand multiparity and reproductive cancer in the Jerusalem Perinatal Study Cohort. Cancer Causes Control. 2016 Feb;27(2):237–47. doi: 10.1007/s10552-015-0701-6. Epub 2015 Dec 15.

9. Lund E. Number of children and death from hormone-dependent cancers. Int J Cancer. 1990 Dec 15;46(6):998–1000. doi: 10.1002/ijc.2910460608.

10. Hinkula M, Kauppila A, Näyhä S, Pukkala E. Cause-specific mortality of grand multiparous women in Finland. Am J Epidemiol. 2006 Feb 15;163(4):367–73. doi: 10.1093/aje/kwj048. Epub 2005 Dec 21.

11. Chan TF, Wu CH, Changchien CC, Yang CY. Mortality from breast, endometrial and ovarian cancers among grand multiparous women in Taiwan. Aust N Z J Obstet Gynaecol. 2011 Dec;51(6):548–52. doi: 10.1111/j.1479-828X.2011.01360.x. Epub 2011 Sep 12.

12. Jaffe DH, Eisenbach Z, Manor O. The effect of parity on cause-specific mortality among married men and women. Matern Child Health J. 2011 Apr;15(3):376–85. doi: 10.1007/s10995-010-0591-x.

13. Lund E. Childbearing in marriage and mortality from breast cancer in Norway. Int J Epidemiol. 1990 Sep ;19(3):527–31. doi: 10.1093/ije/19.3.527.

14. Løchen M-L, Lund E. Childbearing and mortality from cancer of the corpus uteri. Acta Obstet Gynecol Scand. 1997 Apr;76(4):373–7. doi: 10.1111/j.1600-0412.1997.tb07996.x.

15. Lund E. Mortality from ovarian cancer among women with many children. Int J Epidemiol. 1992 Oct;21(5):872–6. doi: 10.1093/ije/21.5.872.

16. Central Bureau of Statistics of Norway.population census 1960. Volume II. Population by sex, age and marital status. Norges offisielle statistikk XII 117, Oslo, 1963.

17. Lettenstrøm, GS. Marriages and number of children – an analysis of fertility trend in Norway, Statistics Norway, No 14, 1965.

18. Mark Schiffman, John Doorbar, Nicolas Wentzensen, Silvia de Sanjosé, Carole Fakhry, Bradley J Monk, Margaret A Stanley, Silvia Franceschi. Carcinogenic human papillomavirus infection. Nat Rev Dis Primers 2016 Dec 1:2:16086. doi: 10.1038/nrdp.2016.86

19. Bevans, R. Akaike Information Criterion | When & How to Use It (Example). Scribbr. Retrieved August 14, 2023, https://www.scribbr.com/statistics/akaike-information-criterion/

20. Bross IDJ. Pertinency of an extraneous variable. J Chron Dis 1967; 20: 487–495.

21. Fedak KM, Bernal A, Capshaw ZA, Gross S. Applying the Bradford Hill criteria in the 21st century: how data integration has changed causal inference in molecular epidemiology. Emerg Themes Epidemiol. 2015 Sep 30;12:14. doi: 10.1186/s12982-015-0037-4.eCollection2015.

22. Husby A, Wohlfahrt J, Øyen N, Melbye M. Pregnancy duration and breast cancer risk. Nature Communications 2018; 9: 4255.

23. Lund E, Rasmussen Busund LT, Thalabard JC. Rethinking the carcinogenesis of breast cancer: The theory of breast cancer as a child deficiency disease or a pseudo semi-allograft. Med Hypotheses. 2018 Nov;120:76–80. doi: 10.1016/j.mehy.2018.08.015. Epub 2018

24. Mellman I, Chen DS, Powles T, Shannon J Turley. The cancer-immunity cycle: Indication, genotype, and immunotype. Immunity. 2023 Oct 10;56(10):2188–2205. doi: 10.1016/j.immuni.2023.09.011.

25. Piccinni MP, Robertson SA, Saito S. Editorial : Adaptive Immunity in Pregnancy.Front Immunol. 2021 Oct 4;12:770242. doi: 10.3389/fimmu.2021.770242.eCollection2021.

26. Yong-Hong Zhang, Ming He, Yan Wang, Ai-Hua Liao. Modulators of the Balance between M1 and M2 Macrophages during Pregnancy. Front Immunol. 2017 Feb 9:8:120. doi: 10.3389/fimmu.2017.00120.eCollection2017

27. Russo J, Balogh GA, Russo IH. Full-term pregnancy induces a specific genomic signature in the human breast. Cancer Epidemiol Biomarkers Prev. 2008 Jan;17(1):51–66. doi: 10.1158/1055-9965.EPI-07-0678.

28. Gillian K Reeves, Sau-Wan Kan, Tim Key, Anne Tjønneland, Anja Olsen, Kim Overvad, Petra H Peeters, Françoise Clavel-Chapelon, Xavier Paoletti, Franco Berrino, Vittorio Krogh, Domenico Palli, Rosario Tumino, Salvatore Panico, Paulo Vineis, Carlos A Gonzalez, Eva Ardanaz, Carmen Martinez, Pilar Amiano, José R Quiros, Maria R Tormo, Kay-Tee Khaw, Antonia Trichopoulou, Theodora Psaltopoulou, Victoria Kalapothaki, Gabriele Nagel, Jenny Chang-Claude, Heiner Boeing, Petra H Lahmann, Elisabet Wirfält, Rudolf Kaaks, Elio Riboli. Breast cancer risk in relation to abortion: Results from the EPIC study. Int J Cancer. 2006 Oct 1;119(7):1741–5. doi: 10.1002/ijc.22001.

29. Guo J, Huang Y, Yang L, Xie Z, Song S, Yin J, Kuang L, Qin W. Association between abortion and breast cancer: an updated systematic review and metaanalysis based on prospective studies. Cancer Causes Control. 2015 Jun;26(6):811–9. doi: 10.1007/s10552-015-0536-1. Epub 2015 Mar 17.

30. Østerbø A. Skammens mødre. Vestlandet forlag 2021

31. Kvåle G, Heuch I, Eide GE. A prospective study of reproductive factors and breast cancer. I. Parity. Am J Epidemiol. 1987 Nov;126(5):831–41. doi: 10.1093/oxfordjournals.aje.a114720.

32. Lambe M, Hsieh CC, Chan HW, Ekbom A, Trichopoulos D, Adami HO. Parity, age at first and last birth, and risk of breast cancer: a population-based study in Sweden. Breast Cancer Res Treat. 1996;38(3):305–11. doi: 10.1007/BF01806150.

33. Collaborative Group on Hormonal Factors in Breast Cancer. Breast cancer and breastfeeding: collaborative reanalysis of individual data from 47 epidemiological studies in 30 countries, including 50302 women with breast cancer and 96973 women without the disease. Lancet. Collaborative Group on Hormonal Factors in Breast Cancer. Lancet. 2002 Jul 20;360(9328):187–95. doi: 10.1016/S0140-6736(02)09454-0.

34. Colditz GA, Rosner B. Cumulative risk of breast cancer to age 70 years according to risk factor status: data from the Nurses’ Health Study. Am J Epidemiol. 2000 Nov 15;152(10):950–64. doi: 10.1093/aje/152.10.950.

35. Ritte R, Tikk K, Lukanova A, Tjønneland A, Olsen A, Overvad K, Dossus L, Fournier A, Clavel-Chapelon F, Grote V, Boeing H, Aleksandrova K, Trichopoulou A, Lagiou P, Trichopoulos D, Palli D, Berrino F, Mattiello A, Tumino R, Sacerdote C, Quirós JR, Buckland G, Molina-Montes E, Chirlaque MD, Ardanaz E, Amiano P, Bueno-de-Mesquita HB, van Gils CH, Peeters PH, Wareham N, Khaw KT, Key TJ, Travis RC, Weiderpass E, Dumeaux V, Lund E, Sund M, Andersson A, Romieu I, Rinaldi S, Vineis P, Merritt MA, Riboli E, Kaaks R. Reproductive factors and risk of hormone receptor positive and negative breast cancer: a cohort study. BMC Cancer. 2013 Dec 9; 13:584. doi: 10.1186/1471-2407-13-584.

36. Lund E, Nakamura A, Snapkov I, Thalabard JC, Olsen KS, Holden L, Holden M. Each pregnancy linearly changes immune gene expression in the blood of healthy women compared with breast cancer patients.Clin Epidemiol. 2018 Aug 6; 10:931–940. doi: 10.2147/CLEP.S163208.eCollection2018.

37. Kezia Gaitskell, Jane Green, Kirstin Pirie, Isobel Barnes, Carol Hermon, Gillian K Reeves, Valerie Beral. Million Women Study Collaborators Histological subtypes of ovarian cancer associated with parity and breastfeeding in the prospective Million Women Study. Int J Cancer. 2018 Jan 15;142(2):281–289. doi: 10.1002/ijc.31063. Epub 2017 Oct 12.

38. Wentzensen N, Poole EM, Trabert B, White E, Arslan AA, Patel AV, Setiawan VW, Visvanathan K, Weiderpass E, Adami HO, Black A, Bernstein L, Brinton LA, Buring J, Butler LM, Chamosa S, Clendenen TV, Dossus L, Fortner R, Gapstur SM, Gaudet MM, Gram IT, Hartge P, Hoffman-Bolton J, Idahl A, Jones M, Kaaks R, Kirsh V, Koh WP, Lacey JV Jr, Lee IM, Lundin E, Merritt MA, Onland-Moret NC, Peters U, Poynter JN, Rinaldi S, Robien K, Rohan T, Sandler DP, Schairer C, Schouten LJ, Sjöholm LK, Sieri S, Swerdlow A, Tjonneland A, Travis R, Trichopoulou A, van den Brandt PA, Wilkens L, Wolk A, Yang HP, Zeleniuch-Jacquotte A, Tworoger SS. Ovarian Cancer Risk Factors by Histologic Subtype: An Analysis From the Ovarian Cancer Cohort Consortium. J Clin Oncol. 2016 Aug 20;34(24):2888–98. doi: 10.1200/JCO.2016.66.8178. Epub 2016 Jun 20.

39. Wu Y, Sun W, Xin X, Wang W, Zhang D. Age at last birth and risk of developing epithelial ovarian cancer: a meta-analysis. Biosci Rep. 2019 Sep 13;39(9):BSR20182035. doi: 10.1042/BSR20182035

40. Fortner RT, Ose J, Merritt MA, Schock H, Tjønneland A, Hansen L, Overvad K, Dossus L, Clavel-Chapelon F, Baglietto L, Boeing H, Trichopoulou A, Benetou V, Lagiou P, Agnoli C, Mattiello A, Masala G, Tumino R, Sacerdote C, Bueno-de-Mesquita HB, Onland-Moret NC, Peeters PH, Weiderpass E, Torhild Gram I, Duell EJ, Larrañaga N, Ardanaz E, Sánchez MJ, Chirlaque MD, Brändstedt J, Idahl A, Lundin E, Khaw KT, Wareham N, Travis RC, Rinaldi S, Romieu I, Gunter MJ, Riboli E, Kaaks R. Reproductive and hormone-related risk factors for epithelial ovarian cancer by histologic pathways, invasiveness and histologic subtypes: Results from the EPIC cohort. Int J Cancer. 2015 Sep 1;137(5):1196–208. doi: 10.1002/ijc.29471. Epub 2015 Feb 26.

41. Jordan SJ, Na R, Weiderpass E, Adami HO, Anderson KE, van den Brandt PA, Brinton LA, Chen C, Cook LS, Doherty JA D. M, Friedenreich CM, Gierach GL, Goodman MT, Krogh V, Levi F, Lu L, Miller AB, McCann SE, Moysich KB, Negri E, Olson SH, Petruzella S, Palmer JR, Parazzini F, Pike MC, Prizment AE, Rebbeck TR, Reynolds P, Ricceri F, Risch HA, Rohan TE, Sacerdote C, Schouten LJ, Serraino D, Setiawan VW, Shu XO, Sponholtz TR, Spurdle AB, Stolzenberg-Solomon RZ, Trabert B, Wentzensen N, Wilkens LR, Wise LA, Yu H, La Vecchia C, De Vivo I, Xu W, Zeleniuch-Jacquotte A, Webb PM. Pregnancy outcomes and risk of endometrial cancer: A pooled analysis of individual participant data in the Epidemiology of Endometrial Cancer Consortium. Int J Cancer. 2021 May 1;148(9):2068–2078. doi: 10.1002/ijc.33360. Epub 2020 Nov 17.

42. Ryoko Katagiri, Motoki Iwasaki, Sarah Krull Abe, Md Rashedul Islam, Md Shafiur Rahman, Eiko Saito, Melissa A Merritt, Ji-Yeob Choi, Aesun Shin, Norie Sawada, Akiko Tamakoshi, Woon-Puay Koh, Ritsu Sakata, Ichiro Tsuji, Jeongseon Kim, Chisato Nagata, Sue K Park, Sun-Seog Kweon, Xiao-Ou Shu, Yu-Tang Gao, Shoichiro Tsugane, Takashi Kimura, Jian-Min Yuan, Seiki Kanemura, Yukai Lu, Yumi Sugawara, Keiko Wada, Min-Ho Shin, Habibul Ahsan, Paolo Boffetta, Kee Seng Chia, Keitaro Matsuo, You-Lin Qiao, Nathaniel Rothman, Wei Zheng, Manami Inoue, Daehee Kang. Reproductive Factors and Endometrial Cancer Risk Among Women. JAMA Netw Open. 2023 Sep 5;6(9):e2332296.doi: 10.1001/jamanetworkopen.2023.32296.

43. Palmer JR, Viscidi E, Troester MA, Hong CC, Schedin P, Bethea TN, Bandera EV, Borges V, McKinnon C, Haiman CA, Lunetta K, Kolonel LN, Rosenberg L, Olshan AF, Ambrosone CB. Parity, lactation, and breast cancer subtypes in African American women: results from the AMBER Consortium. J Natl Cancer Inst. 2014 Sep 15;106(10):dju 237. doi: 10.1093/jnci/dju237. Print 2014 Oct.

44. Fortner RT, Sisti J, Chai B, Collins LC, Rosner B, Hankinson SE, Tamimi RM, Eliassen AH. Parity, breastfeeding, and breast cancer risk by hormone receptor status and molecular phenotype: results from the Nurses’ Health Studies. Breast Cancer Res. 2019 Mar 12;21(1):40. doi: 10.1186/s13058-019-1119-y.

45. Bakken K. Patterns of use and medical consequences of Hormone Replacement Therapy. The Norwegian Women and Cancer study (pages 23-24. Dept. of Pharmacy, Faculty of Medicine, UiT The Arctic University of Norway.

46. Lund E. Pilot study for the evaluation of completeness of reporting to the Cancer Registry. In Incidence of Cancer in Norway 1978. The Cancer Registry of Norway, 1981, Oslo, Norway.

47. Cancer registration in Norway. The incidence of cancer in Norway 1959-1961. The Norwegian Cancer Society 1965, Oslo, Norway.

